# Calling for diversity: improving transfusion safety through high-throughput blood group microarray genotyping

**DOI:** 10.1101/2023.12.15.23299980

**Authors:** Michael Wittig, Tim Alexander Steiert, Hesham ElAbd, Frauke Degenhardt, Luca Valenti, Daniele Prati, Luisa Ronzoni, Luis Bujanda, Jesus M. Banales, Natalia Blay, Pietro Invernizzi, Maria Buti, Agustín Albillos, Javier Fernández, Nicoletta Sacchi, Antonio Julià, Anna Latiano, Rafael de Cid, Mauro D’Amato, Rosanna Asselta, Matthias Laudes, Wolfgang Lieb, David Juhl, Christoph Gassner, Andre Franke

## Abstract

Blood transfusions, conducted between donors compatible in their red blood cell (RBC) antigens, play a life-saving role in transfusion medicine. Genetic differences at blood group loci between ethnicities result in diversity and altered frequency of RBC antigens that need to be considered in blood transfusion. Consequently, comprehensive, and accurate blood group antigen typing is especially relevant for inter-ethnic blood transfusions and for minorities underrepresented in the donor population. Blood group microarray genotyping is a cost-efficient and scalable method for comprehensive blood group typing. Previously, however, microarray typing has been challenging for the clinically important blood group systems Rh and MNS, as these feature highly paralogous genomic loci leading to mixed signals. We here present an approach for accurately typing blood group systems, including Rh and MNS variations, that we benchmarked in an ethnically diverse cohort. We tested its performance using gold-standard, diagnostic-grade MALDI-TOF data from 1,052-samples, including 334 CEPH diversity samples and applied the approach to 4,999 samples of a COVID-19 genetics study. Overall, we obtained a 99.95% benchmarking concordance and 99.65% call rate. In summary, we provide a highly accurate and cost-efficient high-throughput genotyping method for comprehensive blood group analysis that is also suitable for ethnically diverse sample sets.

## INTRODUCTION

Each year, blood transfusions save millions of lives.^1^ Although lifesaving, blood transfusions are not free of risks.^2^ The majority of blood in American blood banks is from white donors of European ancestry, and despite counteracting efforts to recruit more members of ethnic minorities to donate, respective donations are still underrepresented.^3,4^ This can be a problem for all members of ethnic minorities, as the population frequency of certain alleles encoding blood group antigens significantly differs between ethnicities.^5^ For example, the FY^b^ antigen is prevalent in 83% of Caucasians but only in 19% of Asians, which particularly affects Asian minorities transfused in Western countries.^6,7^ In addition, according to Anderson *et al.*,^8^ individuals belonging to ethnic minorities in Western countries have a higher prevalence of blood use than Caucasians which makes them more dependent on blood transfusions and hence more vulnerable.

The presence or absence of blood group antigens radically differ within populations. **Table 1** shows a selected set of antigens of different blood group systems that are defined as clinically significant by the International Society of Blood Transfusion (ISBT)^9^ for individuals of Black African, Caucasian, and Asian descent. Differences in abundance for the selected antigens are obvious, although finer ethnicity-resolved data is still not available for most antigens. In reality, the distribution of blood group phenotypes can even be very heterogeneous among ethnic groups living in proximity,^10^ subgroups within the same ethnicity,^11^ or can be restricted to religious groups^12^ or even single families.^13^

**Table 1:** *Selected antigens of clinical significance and their frequencies in different ethnicities,* according to Reid *et al.*^6^. Abbreviations: Chr.: encoded on chromosome; mod.: moderate; sev.: severe; HTR: hemolytic transfusion reaction; HDFN: hemolytic disease of the fetus and newborn; *: rare occurrence.

| General |  |  | Frequency by ethnicity [%] |  |  | Clinical significance |  |
| --- | --- | --- | --- | --- | --- | --- | --- |
| Blood group system | Chr. | Antigen | Asian | Black | Caucasian | HTR | HDFN |
| ABO (ISBT001) | 9q34 | A | 28 | 27 | 43 | no to sev. | no to mod. |
|  |  | B | 27 | 20 | 9 | no to sev. | no to mod. |
| MNS (ISBT002) | 4q31 | S | no data | 31 | 55 | no to mod.* | no to sev.* |
| Rh (ISBT004) | 1p36 | D | 99 | 92 | 85 | mild to sev. | mild to sev. |
|  |  | C | 93 | 27 | 68 | mild to sev. | mild |
|  |  | c | 47 | 98 | 80 | mild to sev. | mild to sev. |
|  |  | Ce | 92 | 68 | 27 | mild | mild |
| Kell (ISBT006) | 7q34 | K | rare | 2 | 9 | mild to sev. | mild to sev.* |
|  |  | Js<sup>a</sup> | no data | 20 | Rare | no to mod. | mild to sev. |
| Duffy (ISBT008) | 1q23 | Fy<sup>a</sup> | 99 | 10 | 66 | mild to sev.* | mild to sev.* |
|  |  | Fy<sup>b</sup> | 19 | 23 | 83 | mild to sev.* | mild* |
| Kidd (ISBT009) | 18q12 | JK<sup>a</sup> | 72 | 92 | 77 | no to sev. | mild to mod.* |
|  |  | JK<sup>b</sup> | 76 | 49 | 74 | no to sev. | no to mild* |

Blood group antigens have far-reaching or even fatal^14^ consequences stretching beyond hemolytic transfusion reaction (HTR) and hemolytic disease of the fetus and newborn (HDFN) as they have been shown to be linked to other disease^15-18^. As for relatively frequent antigens, allele frequencies of rare antigens often differ among ethnicities, implying prevalence in transfusions beyond ethnicities.^19-22^ For the 360 reported red cell antigens in 45 blood group systems,^23^ the risks of rare antigens can sum up to be reasonably high on the individual level, not even including many unknown and recently discovered^14,19^ antigens from insufficiently studied ethnicities.

Phenotypic differences as well as underrepresentation of ethnic minorities in blood transfusion are long-known and have been addressed but not yet solved. In many Western countries specific recruitment programs have been evaluated to recruit blood donors of diverse backgrounds, however equal representation is still far from being reached.^4,24,25^ Consequently, ethnic minorities in Western countries can be disadvantaged in terms of supply with safe and matching blood units. Even though, life-threatening conditions in the overwhelming majority of cases can be prevented through the sophisticated safety measures of blood banks, the lack of information on non-ABO and non-RhD antigens in many cases leads to a suboptimal distribution of blood units or the waste of precious blood on laborious crossmatch experiments.

The consequences of inter-ethnical donor-recipient incompatibility of blood units can include alloimmunization, which is especially prevalent in chronically transfused members of minorities^7^. One example is sickle cell disease (SCD) that often requires chronic transfusion and has > 80% of its total prevalence in Africa^26^ and hence is more prevalent in people of African descent. Castro *et al.*^27^ determined the quota of phenotype matched blood for individuals of African descent living in the US, to be more than 24-fold higher within African-American cohorts (14.6%) than between Caucasian donors and SCD recipients (0.6%).^27^ The resulting alloimmunization was reported in another study by Olujohungbe *et al.*^28^ where they found that 76% of regularly and locally transfused SDC patients in Britain were alloimmunized, most of them (63%) even with multiple alloantibodies, while this could only be detected in 3% of a Jamaican cohort not constituting an ethnic minority in Jamaica.

Although it is known that extended matching in recipient-donor-matched transfusions can help to reduce the rate of alloimmunization,^29^ in the US and many other developed countries standard typing is still limited to ABO and RhD^30^. Despite in many cases a more comprehensive matching (including MNS, Lewis, Kell, Duffy, and Kidd) for chronically transfused patients is done,^31^ serological test reagents are often unavailable^32^ for many clinically significant antigens other than the most common (ABO, Rh antigens and approximately 18 more). Whereas perfect antigen matching of all transfusions is unrealistic, the current state is insufficient and can be improved.

Cost-effective and high-throughput blood group typing by genotyping could be a possible solution, to insufficient blood type matching, as one could type multiple if not all blood group antigens in a single test for large donor panels. More recently, feasibility for blood group genotyping by whole genome sequencing,^33^ whole exome sequencing, applying next-(NGS),^34^ and third-generation sequencing (TGS)^35^ has been demonstrated. Single nucleotide polymorphism (SNP)-based microarrays are technically appropriate for donor screening since they are highly accurate, highly scalable, widely distributed and feature low per sample cost.^36-38^ However, with SNP microarrays blood group genotyping has not been performed in sufficient accuracy for all clinically important blood group systems.^39^ Inaccuracies in microarray-based testing have previously been described especially for the highly relevant and diverse Rh and MNS systems,^36,40^ specifically the *RHD* and *RHCE* genes *for* Rh, as well as *GYPA*, *GYPB*, *GYPE*/*GYPC* genes in the MNS/Gerbich systems. Due to the paralogous nature of their encoding loci as well as the length limitation of 50 bp (Illumina) of the detection probes, some probes can emit mixed signals tracing back to unspecific binding of fragments from paralogue genomic locations. These inaccuracies made microarray-based typing error-prone, even in homogenous cohorts.^40^

Figure 1 shows the implications of paralogous sequences in microarray genotyping. Sequence paralogs in the targeted area lead to the binding of fragments originating from both genomic loci to the array affixed probes. This results in overlapping, mixed signals that foster inaccuracies for the highly clinically important systems. Such inaccuracies hamper the adoption of the technology, as redundancy of serological and genomic testing cannot be avoided. Reliable phenotyping based on genotyping is also dependent on accurate reference allele information. To ensure this, the ISBT working party on Red Cell Immunogenetics and Blood Group Terminology^23^ documented blood group alleles and their manifested phenotypes. This enables the holistic and reliable phenotype calling based on genotyping data of all blood group antigens at once, even including exceedingly rare antigens, while the accuracy will continue to improve in the future.

**Figure 1:**
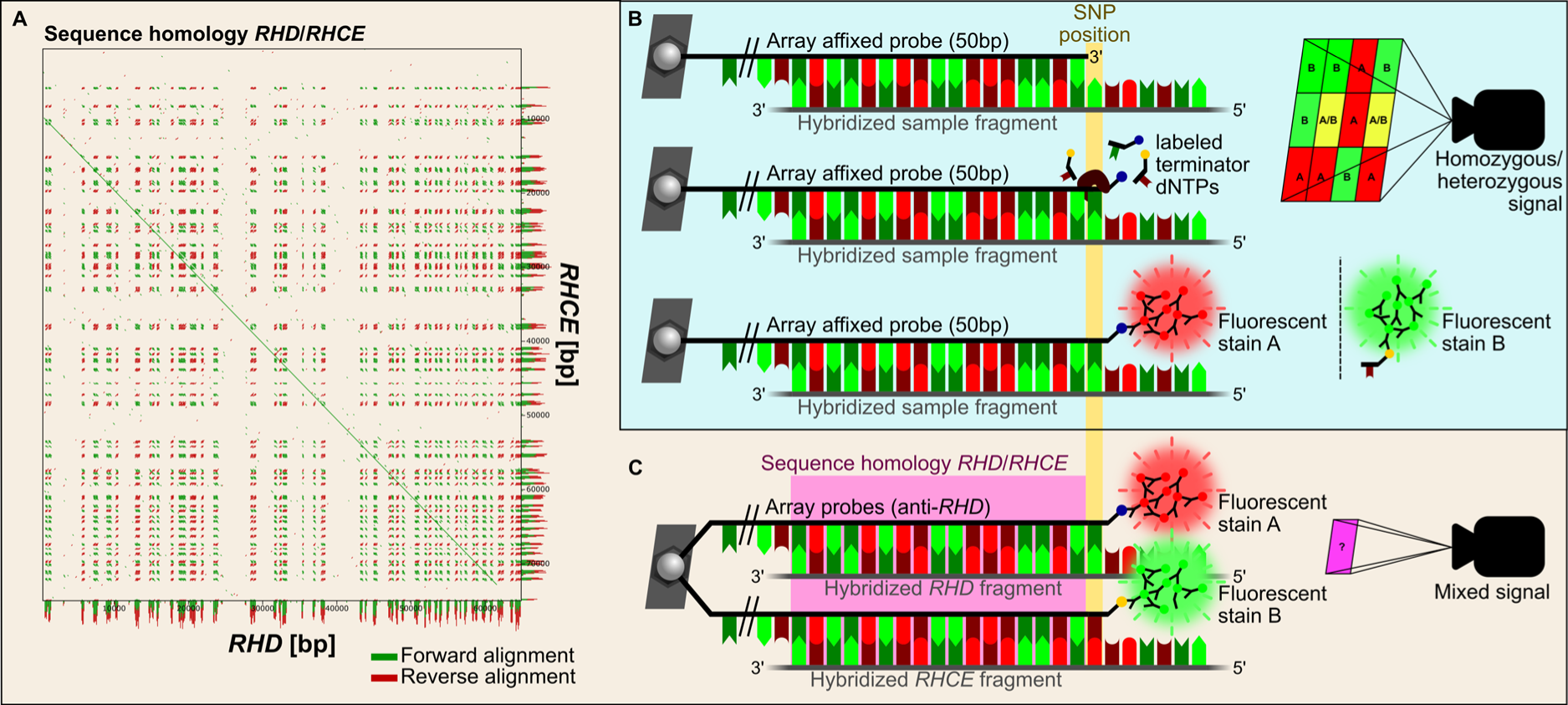
RHD/RHCE paralogy and implications of microarray genotyping for such sequences. (**A**), Dot plot of *RHD* (NG_007494.1, x-axis) and *RHCE* (NG_009208.3, y-axis) ISBT genomic reference sequences created with the *YASS* genomic similarity search tool.^41^ The diagonal line indicates the successful forward alignment and hence the sequence similarity of the two genes over almost their entire sequence. (**B**) Schematic approach of microarray genotyping. An immobilized probe on a bead chip designed against a specific genomic locus is hybridized with a target fragment. A polymerase extends the 3’ end of the probe by one base according to the template sequence that represents the desired SNP position. dNTPs are labeled with a specific hapten for subsequent fluorescent staining. Depending on the respective extended base and the wavelength of the fluorescent signal detected, the genotype for the desired SNP position can be determined for homozygous A or B, as well as heterozygous AB individuals. (**C**) Due to sequence paralogies, such as in *RHD* and *RHCE*, the 50 bp probes are not unique enough in the genomic context to exclusively bind their desired genomic fragments. As a result, signals detected by the camera constitute complex mixed signals from both paralogous loci as well as from different alleles present that cannot be easily interpreted.

We here present and benchmark an approach for reliable single test blood group antigen typing that accommodates comprehensive antigen matching and achieves high phenotype concordance in 14 relevant blood group systems, including Rh and MNS antigens of paralogous loci. While we target and genotype variations across 41 blood group systems, benchmarking is done for only 14, since other systems lack gold standard reference methods to compare against. To simulate the potential use case, we test a 1,052-sample cohort simulating a donor population, 718 German samples, and members of ethnic minorities, 334 CEPH diversity samples.^42^ The comparison involves gold standard serological and MALDI-TOF^43^ typing for ABO antigens and for all remaining antigens, respectively. Compared to blood group antigen typing of the same extent, the consumable costs are significantly more economical, all antigens are typed at once, and the throughput can be easily scaled to several thousands of samples per week. Moreover, we freely share the design of our custom designed genotyping microarray chip targeting 1,112 SNPs associated with blood group variations across 41 blood group systems and provide the analysis scripts and software that utilize deep learning techniques to determine Rh and MNS antigens. Additionally, our database for the association of genotypes with phenotypes is made publicly available (**Supplementary Table 1**).

## METHODS

### Ethical agreement

The use of the German cohort samples was approved by the University Medical Center Schleswig Holstein Institutional Review Board of the Medical Faculty of Kiel University, Kiel Germany in AZ: A103/14 and AZ: 20-1785-101. Informed consent was obtained from all the patients.

### Description of the cohort used in the benchmark study

For the benchmark we used 8 CEPH-1000genomes^44^ DNA samples and 326 DNA samples of the Human Genome Diversity Project (HGDP)-CEPH^45^ panel as provided by CEPH Biobank, Paris, France [BIORESOURCES] as well as a 718 samples from a German cohort used in another study.^46^ The CEPH samples included in this study are composed of individuals originating from the following regions: America: Brazil (n=23), Colombia (n=10), Mexico (n=17); Asia: Cambodia (n=1), China (n=24), Japan (n=8), Pakistan (n=50), Siberia (n=8); Middle East: Israel (Carmel: n=11, Central: n=19, Negev: n=16); North Africa: Algeria (M’zab: n=9); Sub-Saharan Africa: Democratic Republic of Congo (n=12), Central African Republic (n=27), Kenya (n=6), Namibia (n=3), Nigeria (n=14), Senegal (n=13), South Africa (n=5); Oceania: Bougainville (n=7). Europe: Italy (n=13), Russia (n=6), Caucasus (n=6), France (n=14), Orkney Islands (n=4), Ancestries originating from Europe (n=8). All samples were previously typed using MALDI-TOF blood group typing (molecular approach developed by Gassner *et al.*^43^ for 14 blood group systems. In addition to MALDI-TOF data, ABO, reference data was serologically determined for 384 samples where whole blood was available.

### Description of the genotyping microarray

For reliable typing in a diverse cohort, a custom-designed SNP microarray (**Supplementary File 1**) was commissioned at Illumina. The SNP microarray was composed out of 1,112 SNPs that were determined from ISBT reference allele tables as well as from the Erythrogene^44^ blood group gene database to be suitable for allele discrimination. We added 1,500 SNP within the Rh and MNS loci to perform correlation analysis identifying SNPs that were useful in allele discrimination for these loci (see following section). The detailed SNP list (probe design, annotation, cluster definition file) is available as **Supplementary File 2**.

### Determination of allele discriminatory SNPs for paralogous loci

To decrease the complexity of our model and ensure convergence of the neural network given our sample size, we determined the allele discriminating SNPs out of the 1,500 SNPs typed on the array for the paralogous loci and evaluated if the B-allele frequency (BAF) in the Illumina raw genotyping data and genotypes of RhCDE- and MNS-SNPs correlated with the frequency of the reference antigen of our reference data. Correlation based on the coefficient by Pearson and Spearman was calculated in R^47^ using the *cor.test* function from the *stats* package.^48,49^ For each system, the SNPs with the highest absolute value for the correlation coefficient (either Spearman or Pearson) were used.

### Blood group phenotype calling from genotyping data of non-paralogous loci

**Supplementary Table 5** lists all the genomic variations that were used to determine the major alleles in the non-paralogous blood group systems. The software used for non-paralogous blood group system typing is written in C++ and statically translates the genotype into phenotype. A schematic overview of the algorithm is shown in **Supplementary** Figure 1. In brief, genomic variations are called using the GenomeStudio software from Illumina and compared to the variations listed in Supplementary Table 5 to assign a blood type for each blood group. For ABO, blood group typing of blood group A was performed by means of exclusion.

### Blood group phenotype calling from genotyping data of paralogous loci

Calling of paralogous loci *(RHCE*, *GYPA*/*GYPB*/*GYPE*) was performed using a deep-learning-based classification approach using neural networks and the TensorFlow machine learning platform^50^. Architecture [neurons; activation function]: input layer [9; ReLu], hidden layer [6, ReLu], output layer [1; sigmoid]. The weights of the model were optimized using RMSprops and the loss calculated using binary cross-entropy function. 100 epochs were used for training. For further details see GitHub. For each individual, and each paralogous locus, we predicted presence or absence of single antigens and combined the prediction results (*e.g.*, M or N separately to MN).

### Reference model construction and validation

A reference classification model was trained on 80% of randomly selected samples from the benchmark cohort. We defined two sample batches, namely CEPH-diversity and non-CEPH-diversity. We ensured that samples of both batches were representatively present in the training and validation cohort (see source code). The performance of the model was determined on the remaining 20% of the individuals (Table S3.1 of **Supplementary File 3**). The output of each classifier was given as class probabilities between 0 and 1. A cut-off of 0.33 was used to make class assignments (antigen absent: <0.33; failed: 0.33-0.67. present: >0.67).

### Description of cohort and methods used in the COVID-19 study

We applied the methodology for blood group typing to a large cohort in the context of COVID-19. All individuals that were analyzed were recruited by the Severe COVID-19 GWAS study group. Recruitment, ethics, sample processing, and statistical analysis are described in detail in the original publication by Degenhardt *et al.* ^51^ and in the **Online Methods**.

## RESULTS

### Overall blood group typing performance

The concordance rates of the 20 examined antigens in ABO, Rh, and 12 other blood group systems are shown in Figure 2. The benchmark reference data was generated using serological typing for ABO antigens and MALDI-TOF^43^ typing for all remaining antigens. Rh (except D+/-) and MNS antigens featuring paralogous loci were called using a deep-learning-based classifier, while all other antigens used statical logic. Overall, 205 to 1,042 individuals were evaluated, depending on the benchmark reference method used. For paralogous loci only independent validation samples were used for benchmarking. Of note, reductions in benchmarking sample sizes in non-ABO antigens were mainly caused by MALDI-TOF dropouts, as our method achieved a call rate of 99.65% for all antigens and samples. The detailed distribution of blood groups status for all tested individuals is shown in **Supplementary Table 6**. The benchmarking can be traced back in detail in **Supplementary File 4**. In total 16 of 20 antigens yielded perfect concordance, including RhD and Cc antigens. The four imperfectly typed antigens ranged from 98.56% (MN antigen of MNS) to 99.74% (ABO) concordance. The mean concordance of the test based on all antigens combines to 99.95%. To facilitate benchmarking for other studies, we make our phenotype results for many additional CEPH samples available to enable other blood group genotyping benchmarking efforts (**Supplementary File 5**).

**Figure 2:**
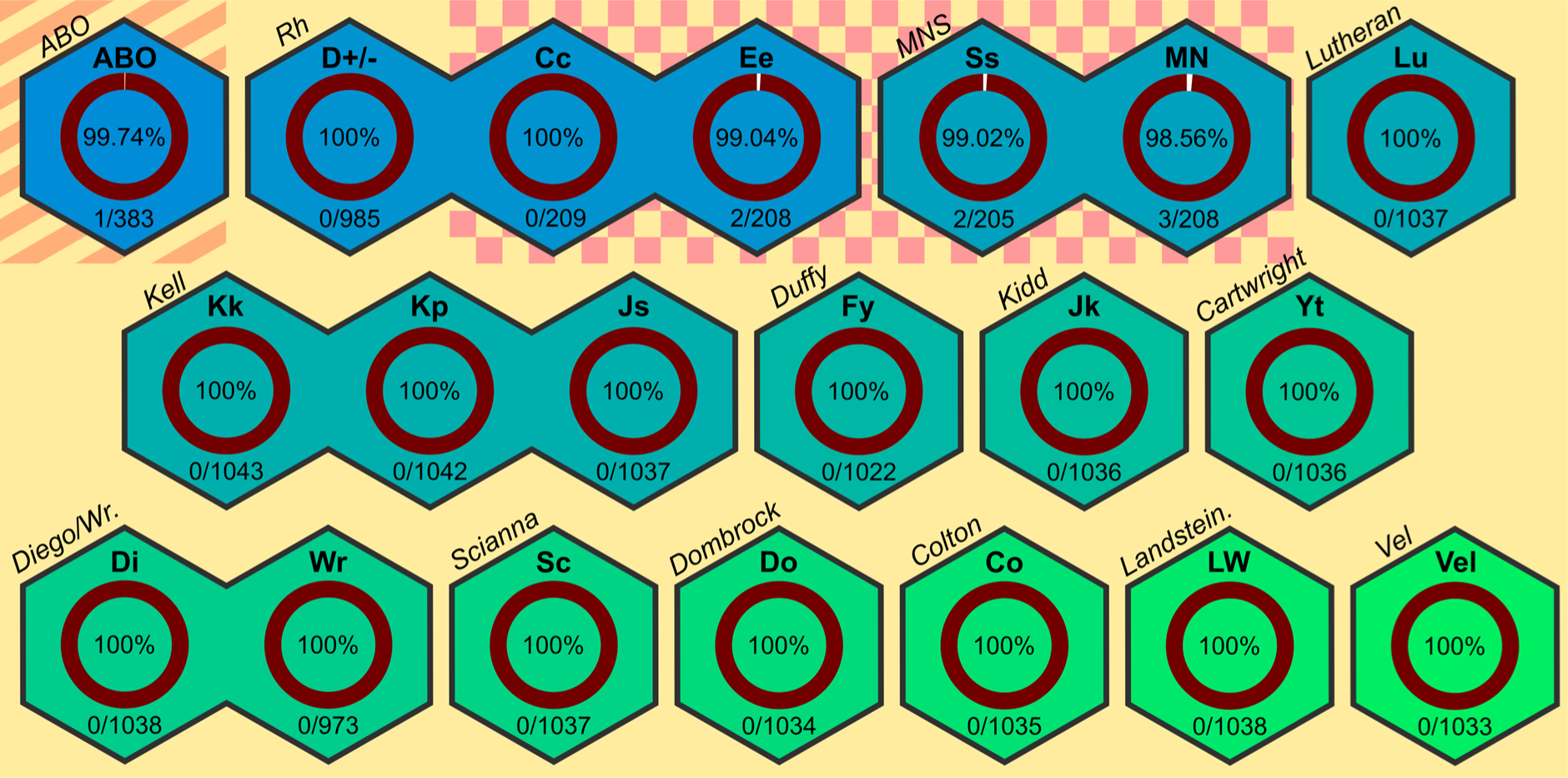
Concordance of SNP microarray-based blood group calling and reference method. Concordance between blood group typing based on microarray genotyping data and respective gold standard reference methods. For ABO (striped background) a serological assay was used as a reference. All other antigens were reference typed with MALDI-TOF genotyping. Using our approach, MNS and some Rh antigens of paralogous loci (plaid background) were called using a deep-learning-based classifier, while the remaining antigens were called using static software logic. The concordance is shown as percentage in the center of the doughnut chart for 20 antigens and blood group systems (upper part of hexagons). Besides ABO and Rh, 12 other blood group systems were typed and benchmarked. For each antigen or system, the number of typing discordances and total benchmark tests is given in the lower part of the hexagons.

For antigens of paralogous loci, we performed additional testing to preclude potential overfitting. To address random allocation of particularly difficult samples in the training set, rather than in the validation set, which could lead to an overestimated accuracy, we performed an iterative re-sampling and re-training evaluation. We re-evaluated the model accuracy based on 18 resampling events with 100 training epochs each, where samples were randomly re-allocated into training and validation set with the only prerequisite that the relative share of diversity panel samples was approximately equal in both, the training and validation set. The resulting model accuracies are depicted in Figure 3. For the c, C and E antigens the developed models yielded perfect accuracy across all re-sampling and re-training iterations. Additionally, this dataset is available in **Supplementary Table 7**.

**Figure 3:**
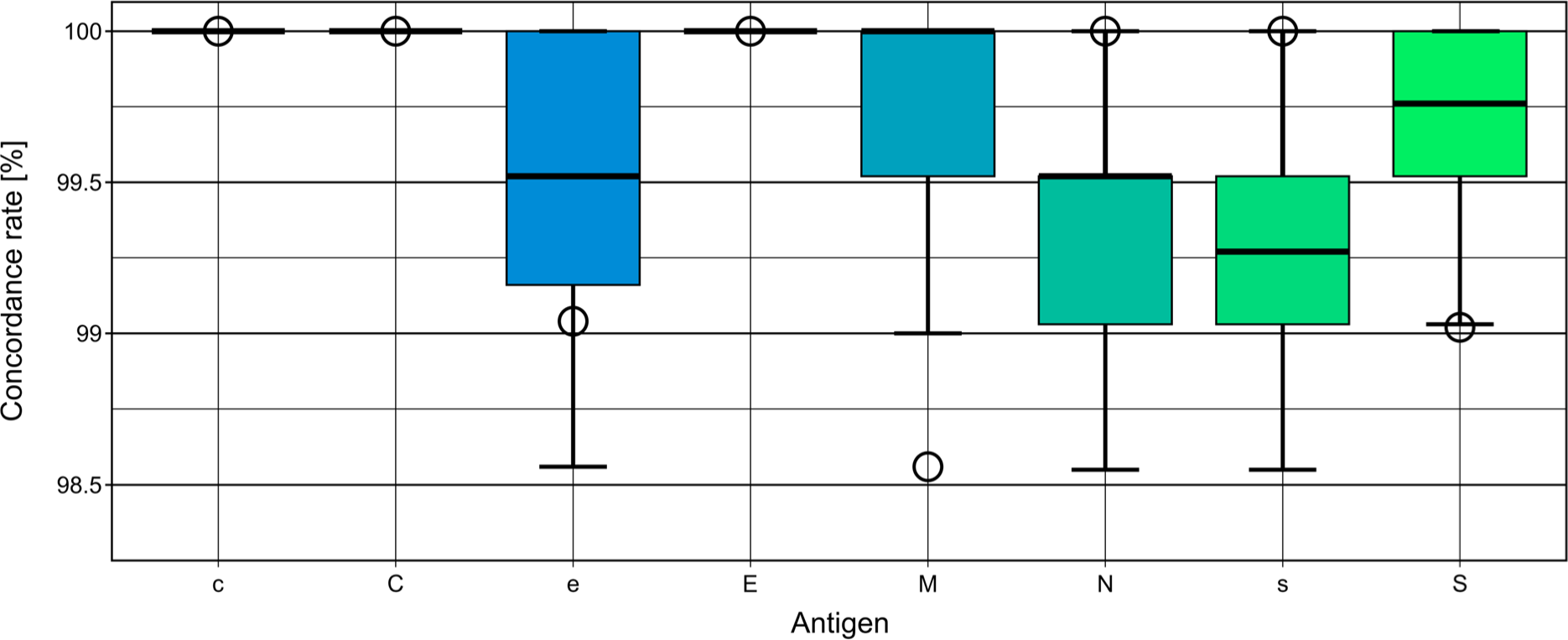
Model concordance rates for the iterative re-sampling and re-training evaluation. The antigens of paralogous loci for the Rh and MNS systems are shown on the x-axis and the distribution of the model concordance rates is shown for all re-sampling and re-training experiments on the y-axis in percent. Lower and upper hinges correspond to 25% and 75% quantile, respectively. Median is indicated by a bold line and lower and upper whiskers represent -1.5 and +1.5 IQR, respectively. The concordance rates of the first model, the one that was used for benchmarking (cf. Figure 2), are shown as empty circles.

Despite the conservative analysis strategy for the non-paralogous blood group loci, that excluded machine learning techniques, the typing accuracy ABO was imperfect with 1 out of 384 samples being not concordant to the reference data. The exact reason for the discrepancy of the serological typing, the ABO reference data comes from serological typing, could not be determined as all relevant genotyping signals were of high quality. Deviations in Rh C, E and MNS antigens are to be expected because of the high diversity of alleles and antigens in the examined multi-ethnic panel and the limited training data. In summary, with a single test performed from DNA, we were able to type 20 blood group antigens from 14 clinically relevant blood group systems in a multi-ethnic and diverse cohort with high accuracy (99.95%). In summary, with our cost-efficient and single test SNP-array-based blood group typing on a comprehensive and still expandable set of blood group systems we achieve accurate results that compare to serological and MALDI-TOF typing.

### Availability of the blood group typing software and data

The developed software for calling blood group phenotypes from genotyping data such as the customized SNP chip used in the study is publicly available at GitHub (https://github.com/ikmb/BloodTypingArray).

### Application of blood group genotyping methodology

As a case study, and to demonstrate the throughput capabilities, we applied our custom blood group typing microarray to a well-characterized cohort of 4,999 COVID-19 patients with respiratory failure from 5 clinical centers in Italy and 6 clinical centers in Spain, previously analyzed in Ellinghaus *et al.*^52^ and Degenhardt *et al.*^51^, **Supplementary Tables 2-5**. In the previous study we identified, for example, a strong association between ABO blood group and susceptibility to SARS-CoV-2. As other blood groups and ABO subtypes were not tested in our original study, we utilized the developed genotyping and analysis framework to perform and to examine the association between other blood groups and SARS-CoV-2.

To determine the statistical significance of genotype frequency differences between COVID-19 patients and the population controls (comparison hereafter referred to as “infection”) as well as different severity levels of COVID-19 in patients only (“severity” analysis), we performed an association analysis. For blood group O (infection; P=1.8x10^-7^; OR=0.65; 95%CI=0.55-0.76) the frequencies of blood types O1O1 and O1O2 was reduced in cases when compared to controls. This indicates that the association observed for blood groups A and O are not subtype-specific in our study. This association was confirmed by other studies.^53-55^ We did not observe any significant associations with COVID-19 in the other 14 analyzed blood group systems neither for the general case-control scenario nor analysis of COVID-19 severity. Although we had limited statistical power to detect weak and moderate associations, our analysis suggests that ABO may be the only relevant blood group system in SARS-CoV-2 susceptibility. No blood group seems to play a significant role in determining COVID-19 severity.

## DISCUSSION

Blood group typing and patient safety in the context of transfusion medicine has constantly evolved over time and improved for the benefit of patients. Although blood transfusions are now safe for most of the population, alloimmunization or even adverse events still occur around the world due to differences in allele frequencies and incompletely matched blood transfusions. Likewise, technologies for genotyping have rapidly evolved and the tools for comprehensive blood group genotyping already exist, waiting to be deployed in blood banking and transfusion medicine.

Technical challenges in especially difficult-to-resolve paralogous genes such as *GYPA*/*GYPB*/*GYPE* and *RHCE*/*RHD* encountered by others have successfully been overcome, for example, by employing the deep-learning-based solution implemented into our typing software. In another study, Gleadall *et al.*^40^ had to determine the copy number state (CNV) of the *RHD* gene to fine tune their analysis for each CNV group but still they observed discordant calls in the Rh system. Even with our limited number of highly diverse HGDP-CEPH panel samples, our developed classifier was able to reliably discriminate antigens encoded in paralogous loci from SNP microarray data. In addition to this, with further training data of samples from diverse ethnicities, our model could still be improved. This will ultimately enable the use of cost-effective and scalable SNP microarray technology for comprehensive blood group genotyping without the need for more costly and less scalable technologies such as NGS or TGS. Using our custom microarray, we here present a benchmark study of 1,052 samples. Moreover, we demonstrated throughput-capabilities by analyzing 4,999 samples of a pan-European cohort in context of COVID-19 susceptibility studies^46,51^. The outcome confirmed previous findings, showing that no additional significant susceptibility blood group loci, besides *ABO,*^54^ seem to exist.

In our benchmark study, the overall accuracy of our methodology with 99.95% is slightly higher than the 99.92% accuracy reported elsewhere for comparable approaches.^40^ This is the case despite involving 14 instead of 9 blood group systems and benchmarking in a more diverse cohort. In our study, the majority of all antigens was typed with 100% accuracy and the lowest concordance in our assay observed for the MN antigen is only 98.56% compared to 99.6% for accurate ABO/RhD diagnostic-grade microplate agglutination assays that are used in clinical practice (^56^ as cited in ^57^). The accuracy of other serological methods in the literature ranges from 85 to 100%,^57^ however, also these studies often do not cover appropriate ethnic diversity or have sufficient sample size.^58,59^ As compared to that, we included 334 diverse CEPH samples in our benchmark study that make up 31.7% of our total cohort. Furthermore, the overall call rate was 99.65% (20,966/21,040 antigen calls) and the dropouts were mainly caused by missing MALDI-TOF benchmark data. We experienced one discordancy in the especially clinically relevant ABO system, potentially leading to a severe HTR. This discordance was further investigated but its root cause could not be unequivocally resolved.

A potential explanation of this ABO mismatch between serological and genotypic testing might be that individual SNP signals resulting from the allele combination represent a variety of possible reference alleles. In this case, the ABO variation 802G>A is representative of the called allele O2,^60,61^ which has a relatively high population frequency, between 5-7%.^62,63^ However, two alternative alleles with this SNP are known, *ABO*AW.08* and *ABO*BW.18*, but are very rare (2/10,000 *cf*. 1000Genomes data^44^). Such ambiguities might be addressed through including additional SNPs, *e.g.*, intronic or flanking, in databases to better distinguish alleles. While unphased microarray data still in very rare cases will result in phenotyping ambiguities, haplotype-phased allele assemblies created with TGS fully resolve the genotypes. The generation of such haplotype assemblies was recently demonstrated, and these will help to improve the quality of the reference allele databases and hence the typing accuracy.^35,64^

To further improve the concordance rates, especially for antigens of the MNS system, we calculated the harmonic mean from the predicted values of both antigens of an antigen pair and used this as a quality score. With a quality score cut-off, it is possible to further improve the accuracy (beyond 99%) for the MNS blood group system, but at minor expense of the call rate. All details about concordance rates, harmonic mean calculation and quality score threshold can be found in **Supplementary File 3**.

In our case study we aimed to demonstrate the high-throughput capabilities of our method and successfully typed 4,999 samples. Further we used the typing results to identify blood group antigens being associated with severe COVID-19. With the ABO system being extensively studied in the context of COVID-19, other blood group systems have been largely neglected or negative results have not been reported. Several studies have suggested that RhD- or O/RhD-(lack of RhD antigen) blood types are protective against COVID-19.^65-67^ In our study, in which we applied the described methodology,^51^ we could not replicate any findings outside of the ABO blood group system, which agrees with others.^54^

As populations become more heterogeneous, since migration increased over the last decades and will continue to do so in the future, there is a growing need for comprehensively matched blood transfusions. This is especially relevant for members of ethnic minorities with chronic disease, requiring RBCs on a regular basis. Our method leveraging SNP microarray genotyping and a deep learning-based software would enable extended blood group matching and as a result make blood transfusion safer. In the beginning, such genotyping methods may be performed to broadly classify patients in need of regular transfusions as well as generating profiles for large numbers of donors. Having comprehensive blood group data for both donors and recipients, optimal pairs can be determined through algorithmic matching. As personalized messaging to donors has been shown to increase repeat donations,^68,69^ back-reporting comprehensive blood group data to donors may also be used to explain why the donor’s blood is especially valuable for people in need of transfusion.

All in all, serological methods will continue to serve as the gold standard method for blood typing but their supplementation through novel genotyping methods will be of high utility where large numbers need to be screened and where analysis time is not critical, *e.g.,* as for donors in blood banks or chronically transfused recipients. Another advantage of genotyping versus serological methods is that it delivers meaningful results even in case of diseases that reduce the levels of blood group antigens.^70^ Additionally, genotyping methods provide an independent platform to resolve forward and reverse typing discrepancies or to aid in other situations.^71^ The examined blood group systems theoretically can be drastically extended for the remaining 26 blood group systems, or even beyond. With the recent advances in chip manufacturing and design, custom microarrays that include tens of thousands of SNPs can be achieved. In our microarray design we already include 1,112 SNPs relevant to blood group genotyping but make only use of 119 (10.7%) SNPs to analyze the 14 blood group systems that we could benchmark with solid reference data. In summary, our comprehensive and cost-effective blood group antigen genotyping method, if used broadly for precision medicine in blood banks, could make transfusion medicine safer, especially for members of ethnic minorities who are most vulnerable.

## Data Availability

The raw genotyping data of the HGDP-CEPH panel are available via the associated database of the Fondation Jean Dausset (www.cephb.fr). All microarray and MALDI-TOF typing results are available as Supplementary Data via medrxiv Online. The data of the German cohort are available upon request from P2N biobank. Access token: P2N_859BH; http://www.uksh.de/p2n/. P2N is a controlled-access human data repository subject to European data protection laws. Therefore, data access is subject to an application, ethics approval by the applicant s ethics board and a data access agreement.

https://github.com/ikmb/BloodTypingArray

## ACKNOWLEDGEMENT

The study was partially funded by a grant from the German Federal Ministry of Education and COVID-19 grant Research (BMBF; ID:01KI20197). The IKMB received infrastructure support from the DFG Excellence Cluster Precision Medicine in Chronic Inflammation (Exc2167), the European Union’s Horizon 2020 research and innovation program European Advanced infraStructure for Innovative Genomics, EASI-Genomics [824110], and the German Research Foundation (DFG) Research Infrastructure as part of the Next Generation Sequencing Competence Network, Competence Center for Genomic Analysis (CCGA) Kiel [423957469].

We thank the Fondation Jean Dausset-CEPH Biobank, Paris, France [BIORESOURCES] for providing and maintaining HGDP-CEPH panel. We acknowledge the technical support in designing the microarrays by Illumina. We further thank Tanja Wesse, Sanaz Sedghpour Sabet, Nicole Braun, Anna Wittmaack, Anja Tanck, and Xiaoli Yi for the excellent laboratory as well as Georg Hemmrich-Stanisak and Teide Boysen for the IT support.

## AUTHORSHIP CONTRIBUTIONS

Contribution: M.W., T.A.S., and A.F. designed the study. M.W. developed the typing software. H.E. provided the deep learning framework. M.W. performed the bioinformatic analysis. F.D. performed the statistical analysis on the COVID-19 cohorts. T.A.S. and M.W. wrote the manuscript with contributions from the other authors. T.A.S. created all figures. L.V., D.P., L.R., L.B., J.M.B., M.D’A., R.d.C., N.B., R.A., P.I., M.B., A.A., J.F., N.S., A.J., A.L., W.L., M.L., and D.J. provided the samples. All authors edited and approved the final manuscript.

## VISUAL ABSTRACT

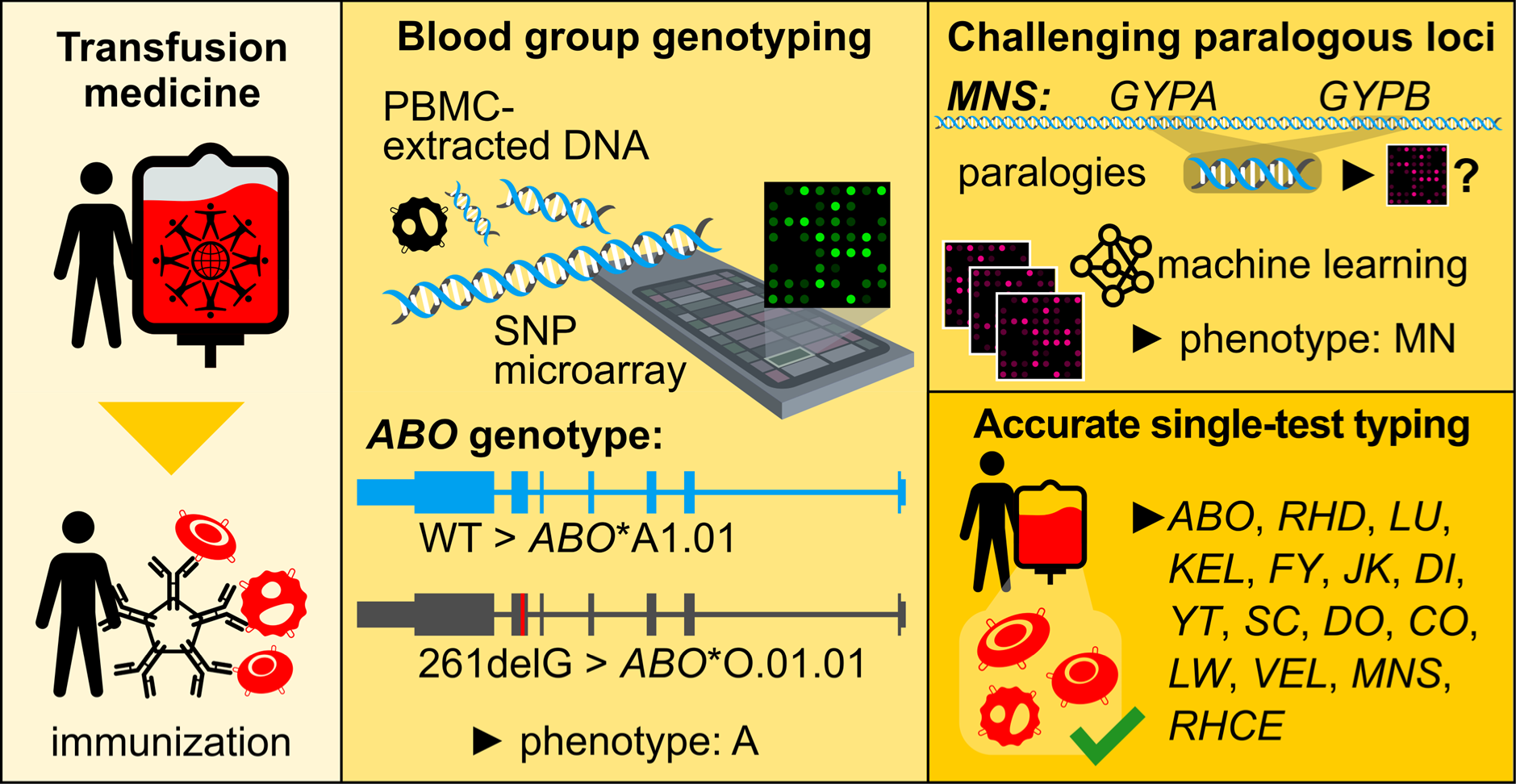

## DATA AVAILABILITY

The software and reference data sets available in the GitHub repository (https://github.com/ikmb/BloodTypingArray).

The raw genotyping data of the HGDP-CEPH panel are available via the associated database of the Fondation Jean Dausset (www.cephb.fr). All microarray and MALDI-TOF typing results are available as Supplementary Data via *medrxiv* Online. The data of the German cohort are available upon request from P2N biobank. Access token: P2N_859BH; http://www.uksh.de/p2n/. P2N is a controlled-access human data repository subject to European data protection laws. Therefore, data access is subject to an application, ethics approval by the applicant’s ethics board and a data access agreement.

## SUPPLEMENTARY DATA

### Supplementary Files

- Online Methods
- Supplementary File 1: Manifest file of the Hemo_MLD_v1_20200225 commissioned at Illumina.
- Supplementary File 2: Detailed complementary files for Hemo_MLD_v1_20200225 commissioned at Illumina.
- Supplementary File 3: Classification of the antigens from paralogous loci
- Supplementary File 4: Concordance check MALDI-TOF vs. microarray data for 334 CEPH diversity samples
- Supplementary File 5: Microarray based molecular blood group typing results for additional CEPH samples

### Supplementary Tables

- Supplementary Table 1: Transcript of the ISBT and Erythrogene databases
- Supplementary Table 2: Frequencies of blood group genotypes in controls in the Italian and Spanish populations.
- Supplementary Table 3: Association analysis of blood group subtypes
- Supplementary Table 4: Association analysis of blood group systems
- Supplementary Table 5: Detailed description of study centers used in the GWAS case study of COVID-19
- Supplementary Table 6: Distribution of the antigens in the benchmarked blood groups
- Supplementary Table 7: Dataset for iterative re-sampling and re-training evaluation

### Supplementary Figures

- Supplementary Figure 1: Schematic overview on the algorithm

---

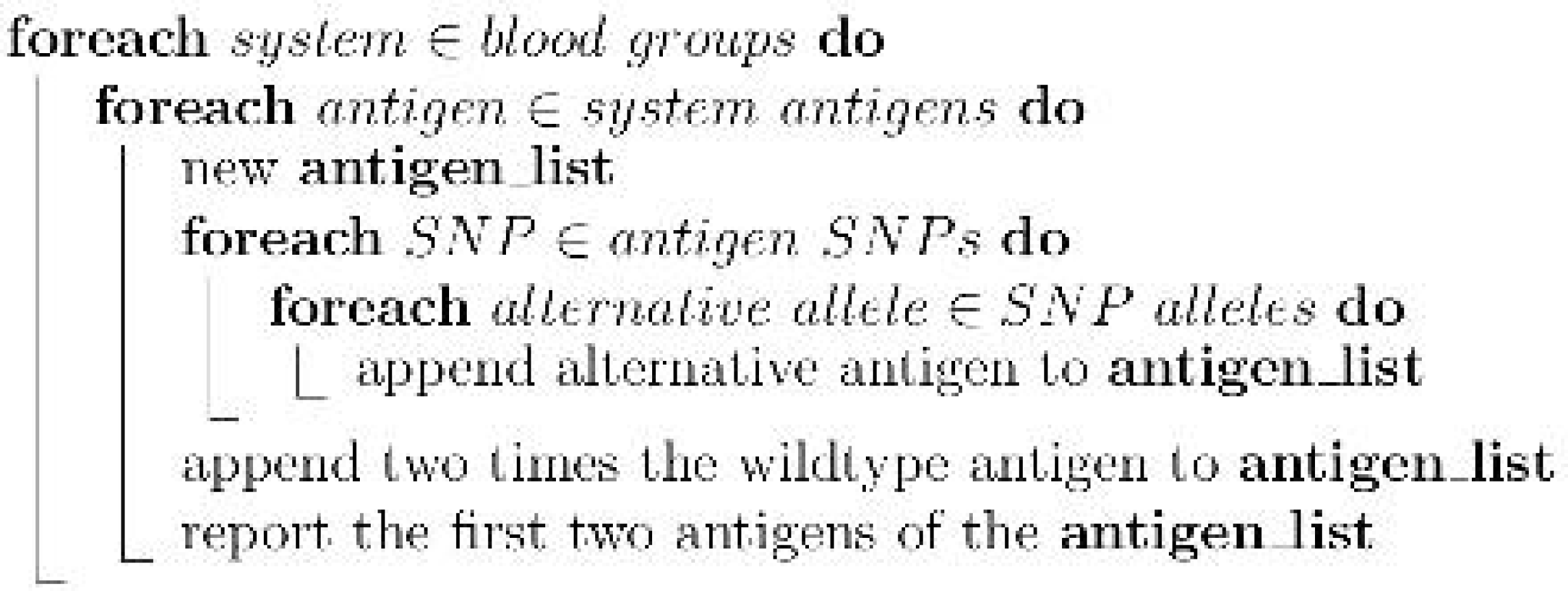

