## Supplementary material for "Calling for diversity: improving transfusion safety through high-throughput blood group microarray genotyping": Online Methods

### DETAILED DESCRIPTION OF COHORT USED IN THE GWAS CASE STUDY OF COVID-19

All individuals that were analyzed in the case study, were recruited by the Severe COVID-19 GWAS study group. Recruitment, ethics, and sample processing are described in detail in the original publication Degenhardt et al[1]. In brief and for the present study, a subset of 4,999 individuals with known age and sex information, 2,229 cases and 2,770 controls, from 11 different study centers across Italy and Spain were analyzed. (**Supplementary Table 5**) Cases were defined as in the original study as individuals with respiratory failure due to a COVID-19 infection. COVID-19 infection was confirmed by a SARS-CoV-2 viral RNA polymerase-chain-reaction (PCR) test from nasopharyngeal swabs or other relevant biologic fluids. Respiratory failure was defined in the simplest possible manner to ensure feasibility: the use of oxygen supplementation or mechanical ventilation, with severity graded according to the maximum respiratory support received at any point during hospitalization (1: supplemental oxygen therapy only, 2: non-invasive ventilatory support, 3: invasive ventilatory support, or 4: extracorporeal membrane oxygenation). Controls were population controls with either unknown or negative COVID-19 status recruited predominantly at blood or bone marrow donation centers. From the 4,999 individuals typed on the blood group typing array, 4,218 remained for analysis after excluding individuals that had failed quality control in the original genome-wide association analysis due to ancestry, relatedness, or genotyping issues[1].

### STATISTICAL ANALYSIS IN GWAS CASE STUDY

A logistic regression analysis was performed in R (vs 3.2.6) separately for individuals from Spain and Italy for different case-control categories as described below. For all analyses the blood type status was coded either as 0 (P=present) or 1 (A=absent). All analyses included age, age2, sex and the interaction between age and sex as well as the first 10 principal components (PCs) calculated from a principal component analysis (PCA) on whole-genome quality-controlled data (full description is given in[1]) as covariates.

#### I. Main analysis

For each individual case-control dataset, COVID-19 patients with respiratory failure (cases, respiratory support status 1-4) were compared with population controls (negative or unknown COVID-19 status). Statistical testing was performed as:

Case/Control ~ blood type status + age + sex + age*age + age*sex + PC1 + PC2 + PC3 +PC4 + PC5 + PC6 + PC7 + PC8 + PC9 + PC10

**II. Severity analysis**

For each individual case dataset, COVID-19 patients were stratified based on their respiratory support status, which we used to define a new case-control status (control = respiratory support status 1; case = respiratory support status 2-4. Statistical testing was performed as:

III. Case only (respiratory support status=1 vs. respiratory support status=2-4) ~ blood type status + age + sex + age*age + age*sex + PC1 + PC2 + PC3 +PC4 + PC5 + PC6 + PC7 + PC8 + PC9 + PC10.

**III. Meta-analysis**

Statistics from the respective analyses in the Spanish and Italian populations were combined using an inverse variance-weighted fixed-effect meta-analysis and the metafor[2] package in R. We conducted statistical analysis only on blood group genotypes with frequencies > 1% or < 99%. P values were corrected using the Holm-Bonferroni method.

**ONLINE METHODS REFERENCES**

1. Degenhardt F, Ellinghaus D, Juzenas S, et al. Detailed stratified GWAS analysis for severe COVID-19 in four European populations. *Hum Mol Genet*. 2022;31(23):3945-3966.

2. Viechtbauer W. Conducting meta-analyses in R with the metafor package. *J Stat Softw*. 2010;36(3):1-48.
