## Supplementary File 3 for "Calling for diversity: improving transfusion safety through high-throughput blood group microarray genotyping"

### 1 Classification of the antigens from paralogous loci

The classification is performed as described in the main manuscript and its performance is shown in table S3.1.

The majority of the values, predicted by the classifier, are close to the extremes 0 and

| System | Antigen | Freq in Training set | Freq in Test Set | Training Size | Test Size | Test Errors | Test failed | Test Precision | Worst Case Success Rate [%] |
|---|---|---|---|---|---|---|---|---|---|
| RhCE | c | 0.8113 | 0.8182 | 832 | 209 | 0 | 0 | 100.00 | 100.00 |
| RhCE | C | 0.5817 | 0.5359 | 832 | 209 | 0 | 0 | 100.00 | |
| RhCE | e | 0.8945 | 0.9187 | 834 | 209 | 2 | 1 | 99.04 | 99.04 |
| RhCE | E | 0.3273 | 0.311 | 834 | 209 | 0 | 0 | 100.00 | |
| MNS | M | 0.7842 | 0.7656 | 834 | 209 | 3 | 0 | 98.56 | 98.65 |
| MNS | N | 0.6583 | 0.6746 | 834 | 209 | 0 | 1 | 100.00 | |
| MNS | s | 0.8382 | 0.8357 | 822 | 207 | 0 | 2 | 100.00 | 99.02 |
| MNS | S | 0.5511 | 0.5604 | 822 | 207 | 2 | 2 | 99.02 | |

**Table S3.1:** Precision for classifying the antigens of RhCE and MNS. These systems are characterized by pseudo SNPs, generated by paralogous genes with highly homologous sequences at the corresponding SNP coordinates. Here we use alternative SNPs from the non homologous regions and trained a tensor flow classifier with more than 800 samples for classifying wether the corresponding antigen is present or not. We show the frequency of the antigen in the training- and test set and the size of both sets. The worst case success rate is calculated, assuming the error rates for the single antigens sum up.

1, but there is also a scatter up to 0.5 (figure S3.1). The closer the value gets to 0.5, the less trustworthy it is. Here you could simply define a range of values in which you define values as untrustworthy. In the graphic, this is shown in red as an example. As

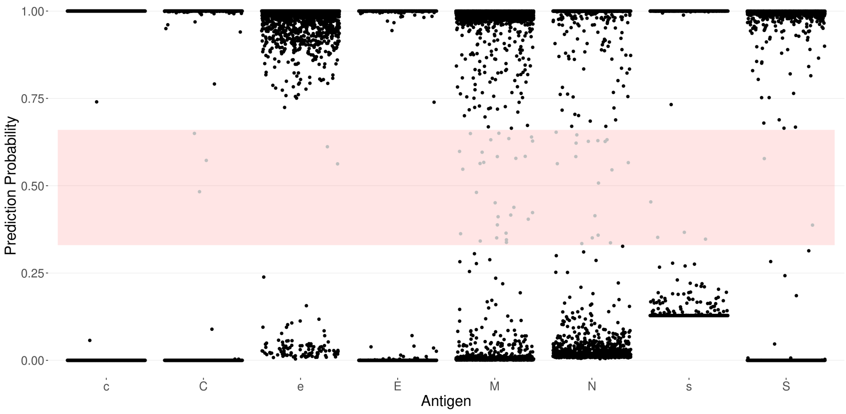


**Figure S3.1:** Distribution of classification probabilities of the different antigens for 5043 samples. Antigens C,c,E,e belong to the RhCE system. The antigens M,N,S,s belong to the MNS system. The red area ranges from 0.33 to 0.66. A prediction value 1 means the classifier is 100% certain, the corresponding antigen is present. A value of 0 means 100% certain it is absent. Values above 0.5 are interpreted by the software as antigen present, below as antigen absent. An area of uncertainty can be defined if only high confident results are wanted (e.g. red area spanning 1/3 of the prediction values).

we evaluate the classification probabilities of each antigen of an antigen pair together, a mean value was deemed as a reliable approximation for a probability score. We have decided to use the harmonic mean, because a good total score can only be achieved if all input values are high enough. The harmonic mean calculation is described here:

$$score = 4 * \frac{ABS(0.5 - X) * ABS(0.5 - Y)}{ABS(0.5 - X) + ABS(0.5 - Y)}$$

where $X$ is the classifier value for the first antigen

where $Y$ is the classifier value for the second antigen

If we now plot the QC scores (harmonic mean) of the true and false predictions separately we see a clear difference in the distribution (figure S3.2). This is just a trend for RHCE, but for MNS there are very different value distributions between true and false. Therefore we set the QC threshold for MNS to 0.8 and for RHCE we refrain from using a QC threshold. If we consider this QC score for the classification the performance for MNS improves with

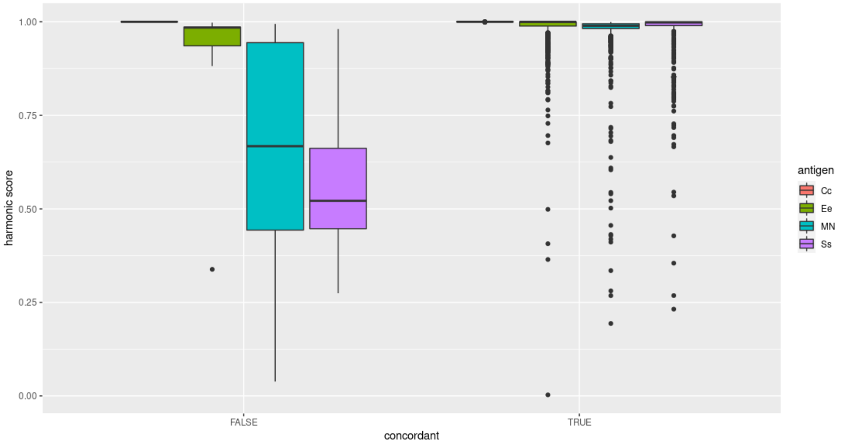


**Figure S3.2:** QC score distribution for the paralogous antigens grouped by concordant/discorcordant. The MNS antigens show a significant difference in the distribution of QC scores between both groups. Applying a 0.8 threshold changes the prediction accuracy as shown in table S3.2. The Cc antigens do not have any discrepancies. The Ee antigens show a significant difference in the distribution, but a very high QC threshold must be applied and many would unnecessarily fail QC.

only 3 % drop outs (table S3.2).

| System | Antigens | Samples | QC Threshold | QC Failed | Errors | Success Rate [%] |
|---|---|---|---|---|---|---|
| RhCE | Cc | 1041 | - | 0 | 0 | 100 |
| RhCE | Ee | 1043 | - | 0 | 7 | 99.3 |
| MNS | MN | 1043 | 0.8 | 36 | 7 | 99.3 |
| MNS | Ss | 1029 | 0.8 | 33 | 2 | 99.78 |

**Table S3.2:** Classifier based antigen prediction with QC threshold. If a quality score is applied the quality of the antigen classification can be improved for the MNS system. Figure S3.2 shows the score distribution for the different antigens and based on these distributions we defined 0.8 for the MNS predictions. While only a small fraction of samples failed QC, we could significantly improve the quality of the antigen prediction (compare with table S3.1 on the preceding page).
